# Metabolic Adaptations to Long-Term Caloric Restriction: Principal Components Analysis of Mass-Spectrometry Metabolomics from the CALERIE™ Phase 2 Trial

**DOI:** 10.64898/2026.02.20.26346654

**Authors:** Melissa C. Orenduff, Emily K. Woolf, Ruiqi Zhang, Daniel W. Belsky, Sai Krupa Das, Waylon Hastings, Justine M. Mucinski, Susan B. Racette, Leanne M. Redman, Reem Waziry, Kari Wong, William E. Kraus, Carl F. Pieper, Kim M. Huffman, CALERIE™

**Affiliations:** Duke Molecular Physiology Institute, Duke University School of Medicine, Durham, NC, USA; Department of Medicine, Duke University School of Medicine, Durham, NC, USA; Reproductive Endocrinology and Women’s Health Laboratory, Pennington Biomedical Research Center, Baton Rouge, LA; Robert N Butler Columbia Aging Center, Columbia University Mailman School of Public Health, New York, NY, USA; Department of Epidemiology, Columbia University Mailman School of Public Health, New York, NY, USA; Jean Mayer USDA Human Nutrition Research Center on Aging at Tufts University, Boston, MA, USA; Department of Nutrition, Texas A&M University, College Station, TX, USA; Institute for Advancing Health through Agriculture, Texas A&M AgriLife Research, College Station, TX, USA; AdventHealth Translational Research Institute, AdventHealth Orlando, Orlando, FL, USA; College of Health Solutions, Arizona State University, Phoenix, AZ, USA; Department of Neurology, Vagelos College of Physicians and Surgeons, Columbia University, New York, USA; Metabolon, Inc. Morrisville, North Carolina, USA; Center on Aging and Development, Biostatistics and Bioinformatics, Duke University, Durham, NC, USA

**Keywords:** CALERIE^TM^, Caloric Restriction, Metabolomics, principal component analysis (PCA)

## Abstract

**Background:** Caloric restriction (CR) improves markers of biological aging, yet long-term effects on the human metabolome remain unclear.

**Objective:** This study examined the effects of CR (2 years) in healthy adults without obesity on circulating metabolites linked to aging and metabolic adaptations.

**Methods:** Untargeted metabolomics was performed using fasted plasma samples collected at baseline, 12, and 24 months (BL, 12M, 24M) from CALERIE™ participants randomized to CR or *ad libitum* (AL) control. A total of 864 known metabolites were identified and grouped into nine biologically coherent super pathways to support pathway-level interpretation (amino acid, peptide, carbohydrate, energy, lipid, nucleotide, cofactors and vitamins, xenobiotics, and partially characterized molecules). Principal component analysis (PCA) summarized metabolite variation, and linear mixed models assessed intervention effects on each PC in group-by-time interactions.

**Results:** Three principal components showed significant group-by-time interactions: PC2 (carbohydrate), PC5 (partially characterized molecules), and PC4 (lipid). Carbohydrate (PC2) and partially characterized metabolites (PC5) decreased from baseline to 12M in both groups; from 12M to 24M, levels stabilized in CR but increased in AL for PC2, while PC5 continued to decline in CR and increased in AL. Lipid metabolites (PC4) decreased in CR and increased in AL at 12M, with the pattern reversing from 12M to 24M. Key contributors included malto-saccharides and related carbohydrate intermediates for PC2, glutamine degradants and lactone sulfates for PC5, and sphingolipids for PC4.

**Conclusion:** This study provided insights into metabolic changes during CR, particularly for carbohydrate and lipid metabolism. Carbohydrate and lipid metabolites that were reduced by CR during the weight loss phase (BL to 12M) followed by stabilization or compensatory responses during the weight maintenance phase (12M to 24M) may link CR-induced changes in metabolism to inflammation. Future research is needed to tease out CR adaptations versus diet related changes in metabolites and explore the functional significance of these metabolic changes during CR for aging and long-term metabolic health.

**Conclusion:** CR produced distinct, time-dependent shifts in carbohydrate and lipid pathways. Early reductions during weight loss followed by stabilization or compensatory responses during weight maintenance suggest dynamic metabolic remodeling that may relate to inflammation-linked mechanisms. Further work is needed to distinguish CR-specific adaptations from dietary influences and to clarify the functional significance of these metabolic changes for aging and long-term metabolic health.

## INTRODUCTION

The Comprehensive Assessment of Long-Term Effects of Reducing Intake of Energy (CALERIE^TM^) study is a landmark investigation into the effects of caloric restriction (CR) on human health and aging. The study aimed to determine the effects of long-term (2 years) CR among healthy adults without obesity compared with an *ad libitum* (AL) control condition. Participants who underwent CR achieved an average of 11.9% caloric reduction and sustained a 10% weight loss over two years (1). Post-hoc molecular analyses suggest modest CR can activate biological pathways associated with healthy aging and potentially delay the onset of age-related diseases (2). Metabolism is a key component in understanding how CR influences health and aging. Previous preclinical studies have demonstrated that CR can lead to significant changes in metabolic processes, including improved insulin sensitivity, reduced inflammation, and enhanced mitochondrial function (3–5). These metabolic adaptations are thought to contribute to the observed lifespan extension and delayed onset of age-related diseases observed in animal models; however, the effects of long-term CR on metabolites and biochemical activities within the ‘healthy’ human metabolome are not well understood.

To address this gap, we performed a global, untargeted metabolomics analysis on circulating metabolites to investigate changes and pathways influenced by CR, which could provide insights into the metabolic mechanisms underlying health benefits and potential impacts on healthy aging.

## METHODS

### CALERIE^TM^ Study Design and Intervention

Detailed CALERIE^TM^ design and methods have been reported previously (6, 7). Briefly, participants were early to mid-life adults [men (20-50 years) and women (20-47 years] and normal weight to overweight (body mass index (BMI) 22.0 to <28.0 kg/m^2^). Participants (n=220) were randomized 2:1 to the CR intervention (n=145) or *ad libitum* (AL) control (n=75) groups (6, 7). Of the 220 participants randomized, 183 individuals (CR = 115; AL = 68) provided plasma samples at all three timepoints [(baseline (BL), 12 months (12M), and 24 months (24M)] and were included in the final analysis. All study participants provided written informed consent, and the study protocol was approved by the Institutional Review Boards at each of the three clinical sites (Washington University School of Medicine, St Louis, MO, USA; Pennington Biomedical Research Center, Baton Rouge, LA, USA; Tufts University, Boston, MA, USA). Duke University (Durham, NC, USA) served as the coordinating site. Phenotypic and clinical data were obtained from the CALERIE™ Biorepository (https://calerie.duke.edu/apply-samples-and-data-analysis). CALERIE^TM^ Phase 2 clinical trial was approved by the Institutional Review Board (IRB:00006471) and registered with clinical trials.gov (<u>NCT00427193</u>) on January 2007.

### Sample Collection and Metabolomic Profiling

Plasma was isolated by centrifugation of blood samples collected in EDTA tubes after an overnight fast (12 h) at three timepoints: BL, 12M, and 24M. Samples were processed and stored at –80°C for analyses at Metabolon® (Durham, NC, USA). Metabolite concentrations were measured using an untargeted Ultrahigh Performance Liquid Chromatography-Tandem Mass Spectroscopy (UPLC-MS/MS) and quantified using the area under the curve of primary MS ions. Of the 1118 untargeted compounds measured, n=864 metabolites had a known identity using Metabolon’s web-service software.

### Analytical Approach

#### Super Pathway Grouping

Metabolites with known identities (n = 1,118) were grouped into nine predefined super pathways to reduce feature complexity and facilitate pathway-level interpretation. These super pathways –– **amino acids**, **peptides**, **carbohydrates**, **energy**, **lipids**, **nucleotides**, **cofactors and vitamins**, **xenobiotics**, and **partially characterized molecules** –– follow Metabolon®, Inc.’s biochemical function–based classification.

#### Principal Component Analysis Within Super Pathways

For each super pathway *SP_j_*, Principal Component Analysis (PCA) was applied to baseline-normalized metabolite concentrations to summarize shared variance among correlated metabolites. Principal components were retained using the elbow criterion, which identifies the point on the scree plot where additional components contribute minimal incremental variance (8, 9). This procedure yielded *X_j_* retained components for each super pathway (**Supplemental Table 1**), providing a reduced set of pathway-level features for downstream modeling.

#### Computation of Participant-Level PCA Scores

To assess change over time, metabolite concentrations for each participant *i* ∈ *N* and metabolite *p* ∈ *P* were normalized at 12M and 24M using baseline statistics:

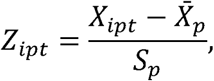

where *X_ipt_* is the concentration of metabolite *p* for participant *i* at time 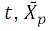, is the mean baseline concentration of metabolite *p*, and *S_p_* is the corresponding baseline standard deviation. Using these normalized values, principal component scores for each retained component *PC_y_* were computed for each participant and timepoint as:

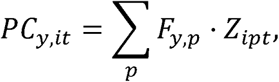

where *F_y,p_* denotes the loading coefficient of metabolite *p* on component *PC_y_*. The resulting time-specific component scores were then used as longitudinal outcomes in mixed-effects models to evaluate the effects of caloric restriction over time.

### Mixed-Model Longitudinal Analyses

To evaluate the effects of group and time on metabolite variation, linear mixed-effects models were implemented in R (version 4.4.0). For each principal component *PC_y_*, the following repeated-measures model was estimated:

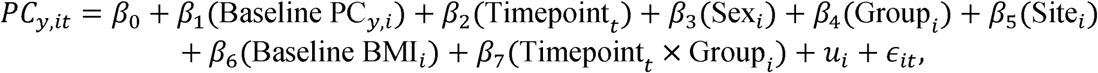

where Timepoint was coded as 0 for 12M and 1 for 24M; Group as 1 for CR and 0 for AL; and Sex as 0 for female and 1 for male. The random intercept u_X_ accounted for within-participant correlation across repeated measures.

The intercept *β*_0_ represents the expected value of *PC_y_* at 12M among females in the AL group, controlling for baseline PC score, study site, and baseline BMI. The interaction term *β*_7_ tests whether the effect of caloric restriction differs between 12M and 24M. Sex, site, and baseline BMI were included as covariates but were not interacted with time or group.

Given the exploratory nature of the analysis, no adjustments were made for multiple comparisons. Results are presented in tabular format, with statistical significance defined as *P* < 0.05.

## RESULTS

### Participant Baseline Characteristics

Study participants (*n*= 183; CR: *n* =115, = AL: *n* = 68) were predominantly female (69%), approximately 38 years old, and White (>70%), with a mean BMI of 25.2□kg/m² (**Table 1**). Although the CR group was prescribed a 25% reduction in caloric intake, the AL group also exhibited slight decreases. Relative to baseline, mean caloric intake declined by 15% in the CR group and 1% in the AL group at 12M, and by 12% and 0.6%, respectively, at 24M (**Table 2**). Body weight in the CR group decreased by 11.7□±□3.8□kg from baseline to 12M and remained stable through 24M (**Table 2**).

**Table 1.** Baseline characteristics for CALERIE^TM^ 2 participants.

|  | ***Ab libitum* (N=68)** | **Calorie Restriction (N=115)** |
| --- | --- | --- |
| **Sex (n(%))** |  |  |
| Female | 47 (69.1%) | 79 (68.7%) |
| Male | 21 (20.9%) | 36 (31.3%) |
| **Age (y)** |  |  |
| Mean (SD) | 38.2 (7.0) | 38.6 (7.3) |
| Median [min, max) | 38.7 [23.2, 50.9] | 40.8 [21.3, 50.9] |
| **Race** |  |  |
| Asian | 3 (4.4%) | 7 (6.1%) |
| Black | 0 (0%) | 0 (0%) |
| White | 50 (73.5%) | 91 (79.1%) |
| Other | 4 (5.9%) | 5 (4.3%) |
| Missing | 11 (16.2%) | 12 (10.4%) |
| **Body Mass Index (kg/m<sup>2</sup>)** |  |  |
| Mean (SD) | 25.2 (1.6) | 25.2 (1.7) |
| Median [min, max] | 24.9 [21.5, 28.4] | 25.1 [21.3, 29.0] |

**Table 2.** Participant characteristics at 12 and 24 months relative to baseline for CALERIE^TM^ 2 participants.

|  | 12 months |  | 24 months |  |
| --- | --- | --- | --- | --- |
|  | *Ab libitum*<br>(N=68) | Calorie<br>Restriction<br>(N=115) | *Ab libitum*<br>(N=68) | Calorie<br>Restriction<br>(N=115) |
| Calorie Restriction (%) | 1.03 (9.08) | 15.10 (7.21) | 0.63 (8.19) | 11.60 (7.09) |
| Fat Free Mass (kg) | 47.90 (8.59) | 46.90 (8.97) | 48.0 (8.38) | 46.80 (9.18) |
| Δ Fat Free Mass | -0.10 (1.11) | -2.05 (1.37) | -0.01 (1.65) | -2.01 (1.77) |
| Fat Mass (kg) | 23.70 (5.95) | 17.30 (3.97) | 24.30 (5.91) | 18.10 (4.17) |
| Δ Fat Mass | -0.26 (2.38) | -6.25 (2.49) | 0.39 (3.34) | -5.41 (2.72) |
| %Δ Weight | -0.69 (4.20) | -11.70 (3.83) | 0.42 (6.01) | -10.40 (4.48) |

Data are presented as mean (SD).

### CR-Mediated Increases in Metabolite Abundance

Across both 12M and 24M, CR was associated with higher concentrations of several metabolite components relative to AL, including Cofactors and Vitamins PC2 (0.46□±□0.12; *P* = 0.0002), Carbohydrate PC2 (0.29□±□0.08; *P* = 0.0003), Carbohydrate PC3 (0.36□±□0.11; P = 0.001) Xenobiotics PC1 (0.45□±□0.18; *P* = 0.02), and Amino Acid PC2 (0.22□±□0.11; *P* = 0.04) (**Figure 1A-E, Supplemental Table 2**). Although the direction of change over time was similar in both groups (e.g., increase, decrease, or plateau), CR consistently maintained higher abundance levels –– except for Carbohydrate PC2 at 24M.

**Figure 1.**
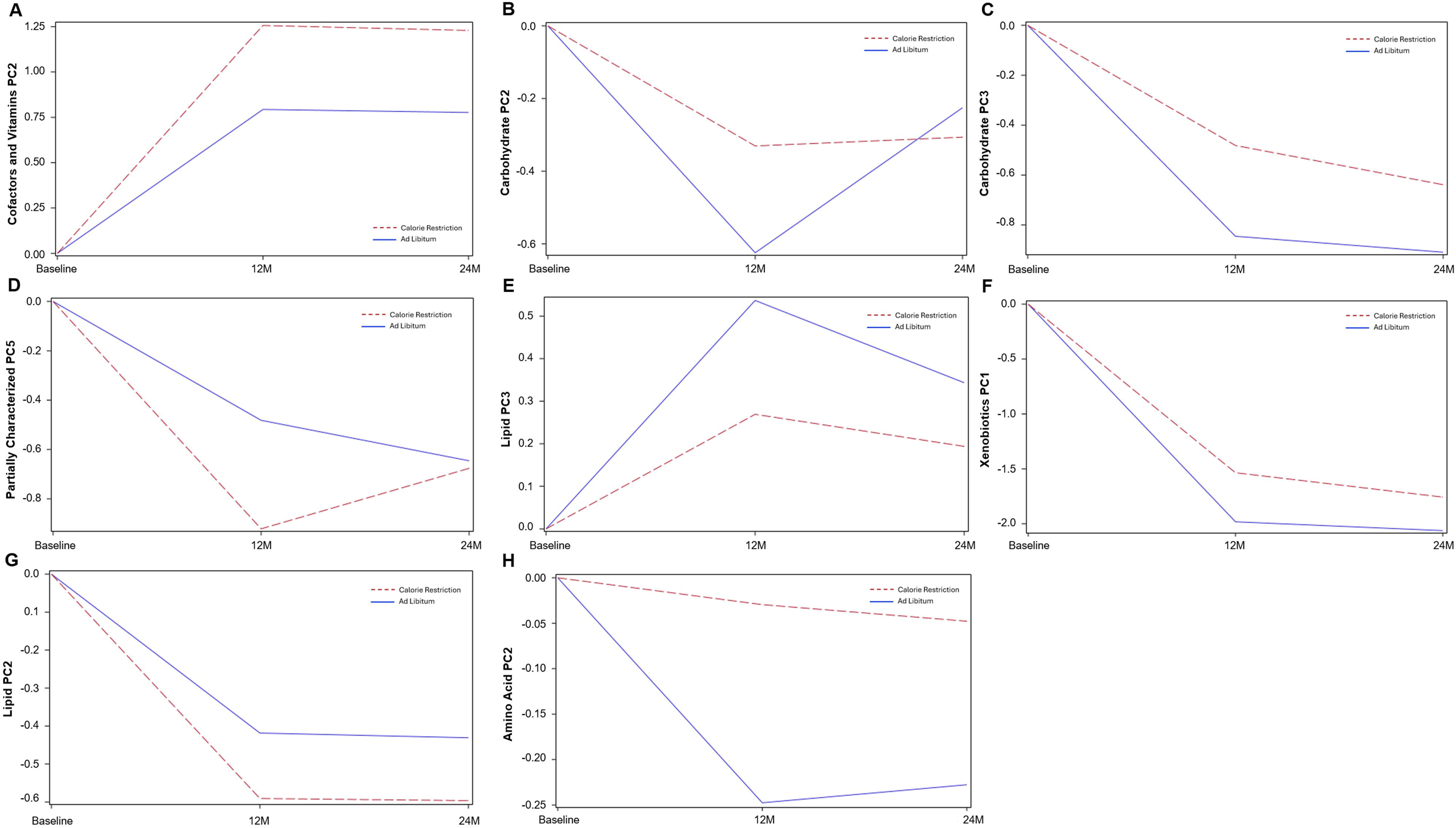
CR⍰mediated differences in metabolite component trajectories over 12M and 24M. Model⍰estimated trajectories of principal component (PC) scores for metabolite groups that differed between CR and AL arms over 12M and 24M. Lines represent predicted mean values from mixed⍰effects models adjusted for baseline metabolite levels, BMI, sex, and site. Statistical estimates and +/-SEs are provided in Supplemental Table 2.

### CR-Mediated Decreases in Metabolite Abundance

Metabolite components that were lower in CR relative to AL across both timepoints included Partially Characterized Molecules PC5 (−0.44□±□0.14; *P* = 0.002), Lipid PC2 (−0.17□±□0.08; *P* = 0.03), and Lipid PC3 (−0.27□±□0.10; *P* = 0.01) (**Figure 1F-H, Supplemental Table 2**). While time-dependent trends were similar across groups, CR consistently exhibited lower abundance levels for these components.

### Time-Dependent Changes in Metabolite Abundance Across Groups

Several metabolites, particularly those related to nucleotide metabolism, exhibited time-sensitive fluctuations independent of intervention group status. Nucleotide PC1 and PC2 decreased at 12M relative to baseline (−1.74□±□0.72, *P* = 0.02 and −1.82□±□0.77, *P* = 0.02, respectively, **Figure 2A-B, Supplemental Table 2**). Between 12M and 24M, Carbohydrate PC2 (0.40□±□0.087, *P* < 0.001) and Peptide PC1 (0.22□±□0.098, *P* = 0.024) (**Figure 2C-D, Supplemental Table 2**) increased, while Cofactors and Vitamins PC1 decreased (−0.19□±□0.097, *P* = 0.047, **Figure 2E, Supplemental Table 2**).

**Figure 2.**
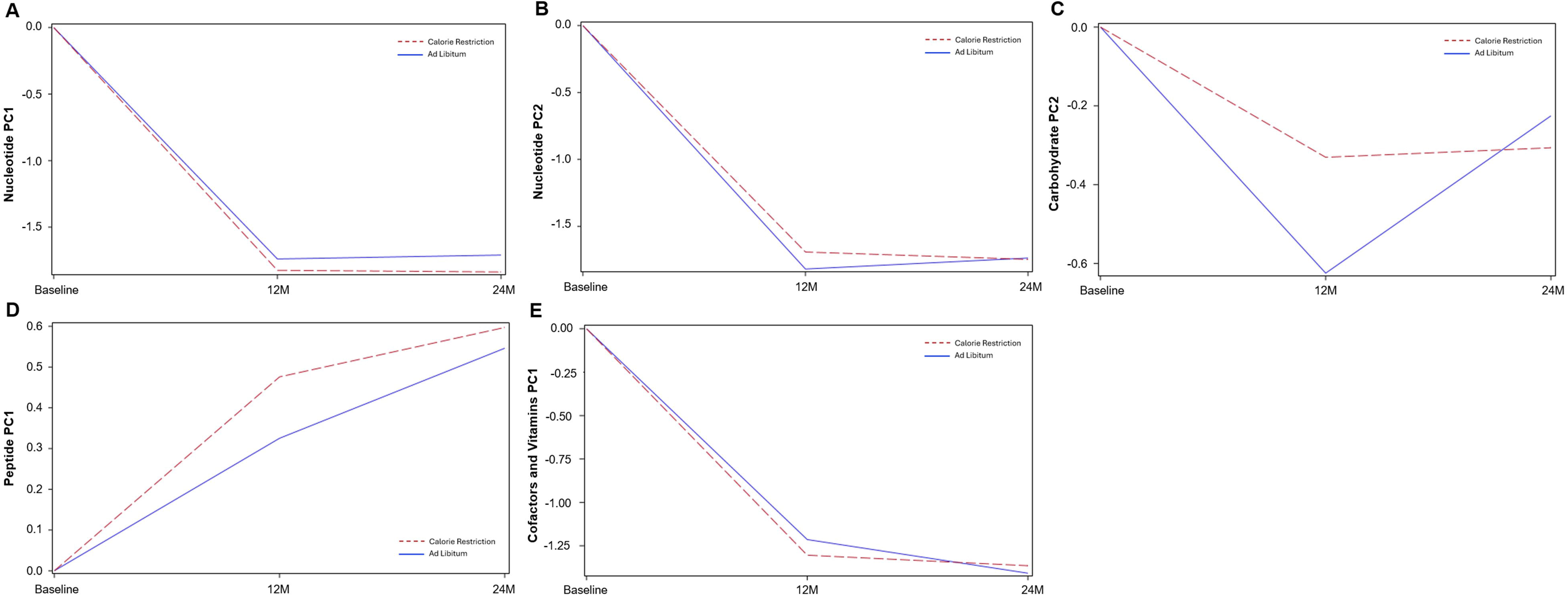
Time⍰dependent changes in metabolite component trajectories independent of intervention group. Model⍰estimated trajectories of principal component (PC) scores for metabolite groups exhibiting ⍰dependent changes between BL, 12M, and 24M, independent of group assignment (CR / AL). Lines represent predicted mean values from mixed⍰seffects models adjusted for baseline metabolite levels, BMI, sex, and site. Statistical estimates and +/-SEs are provided in Supplemental Table 2.

### Time-Dependent Effects of CR on Metabolite Abundance

Significant time□×□group interactions were observed for Carbohydrate PC2, Partially Characterized Molecules PC5, and Lipid PC4 (**Figure 3A-C, Supplemental table 2**). For Carbohydrate PC2, the magnitude of the CR vs. AL group effect decreased by 0.37□±□0.11 at 24M relative to 12M (*P* = 0.0007, **Figure 3A, Supplemental table 2**); both groups showed reductions from baseline to 12M, but from 12M to 24M, levels plateaued in CR and increased in AL. For Partially Characterized Molecules PC5, the group effect increased by 0.41□±□0.15 at 24M compared to 12M (*P* = 0.009, **Figure 3B, Supplemental table 2**); both groups showed initial declines, but CR exhibited a rebound from 12M to 24M, while AL continued to decline. Lipid PC4 showed divergent patterns: the CR group decreased from baseline to 12M and then increased above baseline by 24M, whereas the AL group showed a slight increase at 12M followed by a decline at 24M. The group effect at 24M was 0.47□±□0.23 units greater than at 12M (*P =* 0.04, **Figure 3C, Supplemental table 2**).

**Figure 3.**
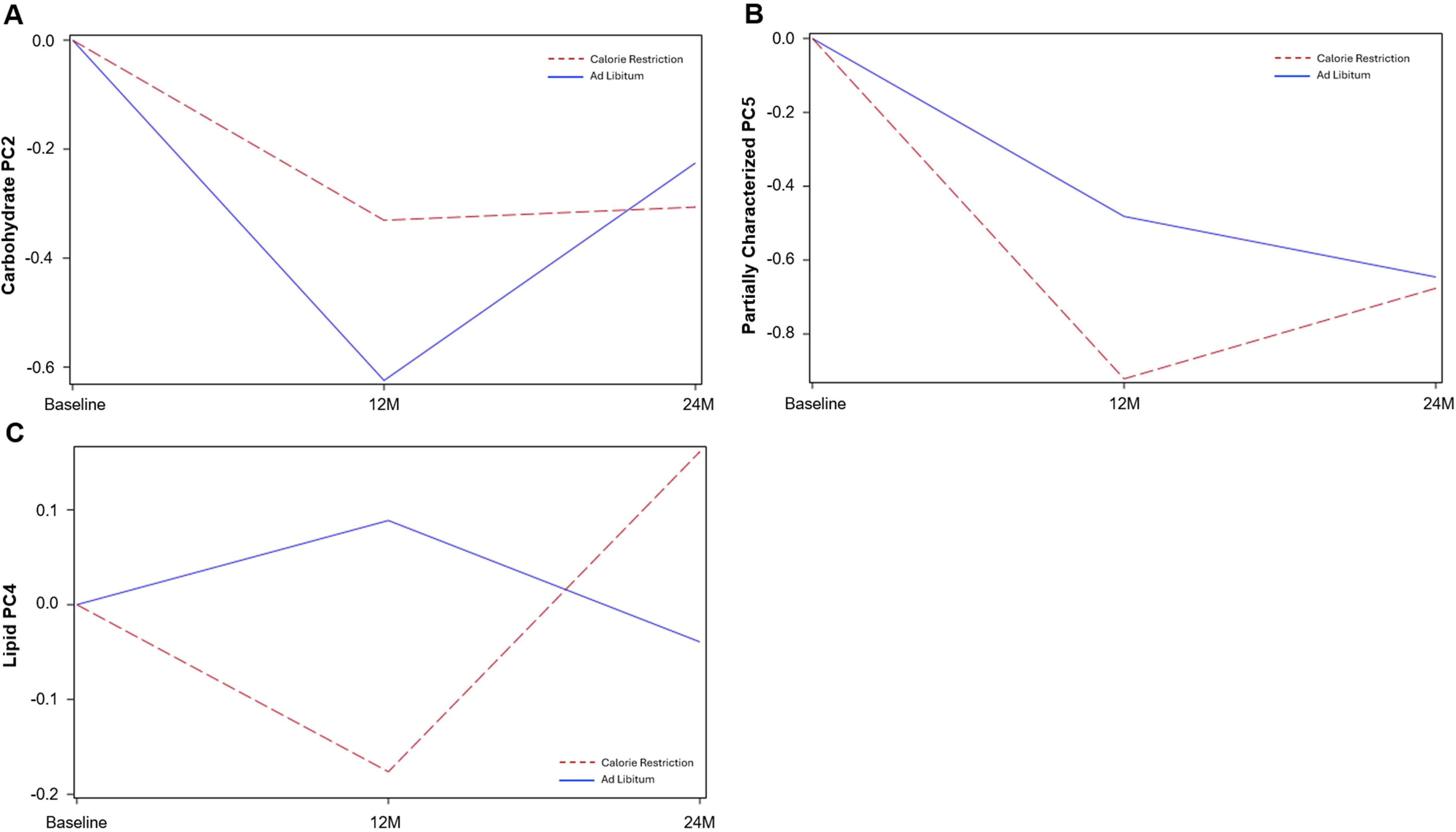
Time⍰dependent effects of caloric restriction on metabolite component trajectories. Model⍰estimated trajectories of principal component (PC) scores for metabolite groups demonstrating significant time□×□group interactions over 12M and 24M. Lines represent predicted mean values from mixed⍰effects models adjusted for baseline metabolite levels, BMI, sex, and site. Statistical estimates and +/-SEs are provided in Supplemental Table 2.

### Principal Component Constituents

Metabolite contributions with the greatest loadings (>2%) on each principal component associated with group, time, and time□×□group effects are listed in **Supplemental Table 3**.

## DISCUSSION

This study provides the first report of how long⍰term CR influences circulating metabolic intermediates over a two⍰year randomized controlled trial in healthy adults without obesity. Using PCA⍰derived metabolite groups and longitudinal mixed⍰effects models, we identified metabolite patterns that changed with CR and differed from AL, revealing both metabolic responses during active weight loss (BL to 12M) and later adaptations during weight maintenance (12M to 24M). These findings highlight metabolic pathways through which CR may exert cardiometabolic and anti inflammatory effects.

### Carbohydrate Metabolism (Carbohydrate PC2)

Carbohydrate PC2 included metabolites such as maltotriose, maltose, maltotetraose, X3 phosphoglycerate, xylose, arabinose, and N⍰acetylneuraminate –– metabolites spanning glycolysis, glycogen breakdown, pentose metabolism, and immune related glycan biology. Maltose⍰ and maltotriose⍰ containing oligosaccharides are classical products of α⍰amylase–mediated starch degradation (10–12). X3⍰phosphoglycerate is a well⍰established intermediate in glycolysis and gluconeogenesis and reflects central energy metabolism (13–15), while xylose and arabinose are pentose sugars typically derived from the breakdown of plant⍰based polysaccharides, including dietary fibers (13, 16, 17); their circulating levels may be influenced by dietary intake or gut microbial metabolism (18, 19), although the specific source cannot be determined from plasma metabolomics alone. N⍰acetylneuraminate (sialic acid) is a terminal glycan with established roles in immune activation, leukocyte signaling, and inflammatory responses (20–22).

Both CR and AL groups showed reductions in these metabolites from baseline to 12 months, but the rebound from 12 to 24 months was markedly attenuated in CR. These patterns likely reflect the initial reduction in energy intake during the first year of CR, followed by longer⍰term physiological adjustments as participants transitioned to weight maintenance. Such adaptations are consistent with prior evidence that sustained caloric restriction induces coordinated metabolic and physiological changes, including reductions in energy expenditure and shifts in metabolic processes that enhance metabolic efficiency and reduce oxidative stress (23). Reduced availability of carbohydrate precursors (e.g., starch⍰sderived saccharides) may also contribute to the observed declines. Notably, sustained reductions in N⍰sacetylneuraminate with CR may reflect anti⍰inflammatory adaptations, given the central role of sialic acids in immune activation and inflammatory signaling (20, 21). Although AL participants exhibited a rebound in these metabolites, the stabilization in CR suggests more durable remodeling of carbohydrate and glycan linked pathways during long⍰sterm CR

### Lipid Metabolism (Lipid PC4)

Lipid PC4 was dominated by sphingolipids, including sphinganine, sphingosine, and their phosphorylated forms –– bioactive lipids central to membrane organization, cell signaling, and inflammatory regulation (24–26). CR produced a sharp decline in these metabolites at 12 months, followed by an increase at 24 months to levels exceeding those of AL. AL showed the opposite pattern, with a modest rise at 12 months and a decline at 24 months. The early decline with CR may reflect acute reductions in inflammatory tone and shifts in sphingolipid⍰mediated signaling during active weight loss, consistent with known CR⍰associated improvements in inflammatory lipid profiles (23, 27). The subsequent rise from 12 to 24 months may represent longer term metabolic adaptation as sphingolipid pools re equilibrate under sustained CR, aligning with evidence that sphingolipid metabolism responds dynamically to changes in energetic state (28, 29). Preclinical studies similarly report CR⍰induced increases in sphinganine species, interpreted as adaptive remodeling of lipid signaling pathways (30, 31). Thus, the biphasic pattern observed here may reflect coordinated, phase⍰specific remodeling of sphingolipid biology across the transition from active weight loss to long⍰sterm CR maintenance.

### Partially Characterized Molecules (PC5)

PC5 included glutamine degradants, pentose acids, branched⍰schain fatty acids, metabolonic lactone sulfates, and glycine conjugates –– metabolite classes linked to amino acid turnover, carbohydrate oxidation, lipid remodeling, and detoxification pathways. CR produced larger reductions at 12 months than AL, followed by partial rebound at 24 months, whereas AL showed progressive declines across both timepoints. Metabolonic lactone sulfate, a xenobiotic⍰srelated metabolite associated with adiposity and cardiometabolic risk (32), declined substantially with CR during active weight loss, consistent with improvements in metabolic health. Glutamine degradants and pentose acids may reflect shifts in amino acid metabolism and reduced inflammatory activity, given that glutamine serves as a key fuel for proliferating immune and tumor cells (33, 34). Reductions in glycine conjugates may indicate lower detoxification demand or changes in dietary patterns toward less processed foods, as glycine conjugation is a major pathway for clearance of xenobiotics and lipid derived acids (35). Because many of these metabolites remain partially characterized, their biological roles require further investigation. Nonetheless, the observed patterns suggest that CR influences multiple interconnected metabolic pathways related to inflammation, amino acid turnover, and detoxification, consistent with broader evidence that CR induces coordinated remodeling across metabolic networks (23, 36, 37).

### Strengths

The randomized controlled design with a well⍰defined *ad libitum* comparator provides a rigorous framework for isolating the metabolic effects of sustained CR. The two⍰syear intervention with fasting plasma collected at BL, 12M, and 24M offers rare longitudinal resolution to distinguish weight⍰loss–related changes at 12M from later weight⍰maintenance adaptations at 24M. Use of a comprehensive untargeted metabolomics platform (864 metabolites across nine super pathways), combined with principal component analysis and mixed⍰effects modeling, enabled systems⍰level characterization of CR⍰related metabolic remodeling while accounting for within⍰person change. The focus on healthy adults without obesity minimized confounding by metabolic disease, strengthening inference about CR specific physiological adaptations. Together, these features make this one of the most detailed longitudinal metabolomic evaluations of CR in humans.

### Limitations

The cohort’s demographic homogeneity (predominantly White, healthy, non⍰obese adults) limits generalizability. Untargeted metabolomics provides relative rather than absolute quantification, and many partially characterized metabolites have uncertain identities or functions, constraining mechanistic interpretation. PCA aggregates metabolites across pathways, potentially reducing biological specificity. Dietary composition was not controlled, making it difficult to separate CR⍰specific effects from changes in food quality or macronutrient intake, and early weight⍰loss effects may overlap with longer term CR mechanisms. Finally, the design did not capture acute metabolic responses occurring more acutely after initiating CR, which may contribute additional insight into early metabolic dynamics.

## CONCLUSION

Sustained caloric restriction drives coordinated, time⍰dependent remodeling across multiple metabolic pathways, with early shifts reflecting active weight loss and later patterns indicating durable physiological adaptation. Across diverse metabolite classes, CR produced consistent divergence from AL intake, revealing a broad reorganization of metabolic networks rather than isolated pathway effects. These findings position CR as a potent modulator of systemic metabolism in healthy adults and underscore the value of longitudinal metabolomics for defining the temporal architecture of human metabolic adaptation.

## Disclaimers

The content is the sole responsibility of the authors and does not necessarily represent the official views of their institutions, the NIA or the USDA.

## Data Availability

All data produced in the present study are available upon reasonable request to the authors

https://calerie.duke.edu/apply-samples-and-data-analysis

https://agingresearchbiobank.nia.nih.gov/studies/calerie/

## ACKNOWLEDGMENTS

We would like to thank the participants of CALERIE^TM^, as well as Metabolon Inc., for the processing and analyses of plasma metabolites.

## AUTHOR CONTRIBUTIONS

MCO coordinated data processing, contributed to statistical modeling and interpretation of results, and led manuscript writing. EKO assisted with metabolite annotation and pathway interpretation. MCO and EKW co led manuscript drafting and revisions. RZ supported data management, quality control, and conducted statistical and bioinformatic analyses under the supervision of CFP. CFP conceived the metabolomics analysis plan and provided statistical guidance. KMH and CFP contributed to study design and to mixed model development and interpretation. DWB, SKD, WH, SBR, LMR, RW, WEK, CFP, KMH contributed to critical review and editing of the manuscript. All authors reviewed and approved the final manuscript.

---

**Supplemental Table 1.** PCA was performed separately within each super⍰pathway, and components were retained based on the scree⍰plot elbow and interpretability criteria. This procedure yielded a specific number of retained components for each super⍰pathway, creating a streamlined set of pathway⍰level features for downstream modeling.

| **Super Pathways** | **No. of metabolites** | **Retained PCs** |
| --- | --- | --- |
| Amino Acid | 215 | 2 |
| Peptides | 36 | 2 |
| Carbohydrates | 26 | 4 |
| Energy | 9 | 2 |
| Lipids | 303 | 4 |
| Nucleotides | 36 | 2 |
| Cofactors and Vitamins | 40 | 4 |
| Xenobiotics | 159 | 5 |
| Partially Characterized Molecules | 15 | 5 |

**Supplementary Table 2.** Differences in significant Superpathway Principal Components (PC) between calorie restriction (CR) and *ad libitum* (AL).

| **Superpathway** | **PC #** | **CR effect +/- SE**<br>**(Factor (group))** | **P-value** |
| --- | --- | --- | --- |
| Cofactors and Vitamins | 2 | 0.46 ± 0.12 | 0.0002 |
| Carbohydrate | 2 | 0.29 ± 0.08 | 0.0003 |
| Carbohydrate | 3 | 0.36 ± 0.11 | 0.001 |
| Partially Characterized | 5 | -0.44 ± 0.14 | 0.002 |
| Lipid | 3 | -0.27 ± 0.10 | 0.01 |
| Xenobiotics | 1 | 0.45 ± 0.18 | 0.02 |
| Lipid | 2 | -0.17 ± 0.08 | 0.03 |
| Amino Acid | 2 | 0.22 ± 0.11 | 0.04 |

***12M and 24M differences in Superpathway PC across CR and AL***
| **Superpathway** | **PC** | **12M effect +/- SE** | **P-value** |
| --- | --- | --- | --- |
| Nucleotide | 1 | -1.74 ± 0.72 | 0.02 |
| Nucleotide | 2 | -1.82 ± 0.77 | 0.02 |
| **Superpathway** | **PC** | **24M effect +/- SE** | **P-value** |
| Carbohydrate | 2 | 0.40 ± 0.087 | 0.000006 |
| Peptide | 1 | 0.22 ± 0.098 | 0.024 |
| Cofactors and Vitamins | 1 | -0.19 ± 0.097 | 0.047 |

***CR Group by time differences in Superpathway PC***
| **Superpathway** | **PC** | **CR by time effect +/- SE**<br>**CR group) x time** | **P-value** |
| --- | --- | --- | --- |
| Carbohydrate | 2 | -0.37 ± 0.11 | 0.0007 |
| Partially Characterized | 5 | 0.41 ± 0.15 | 0.009 |
| Lipid | 4 | 0.47 ± 0.23 | 0.04 |

Effects shown are parameter and standard error (SE) when controlling for baseline metabolite levels, BMI, sex, and site.

**Supplemental Table 3.**
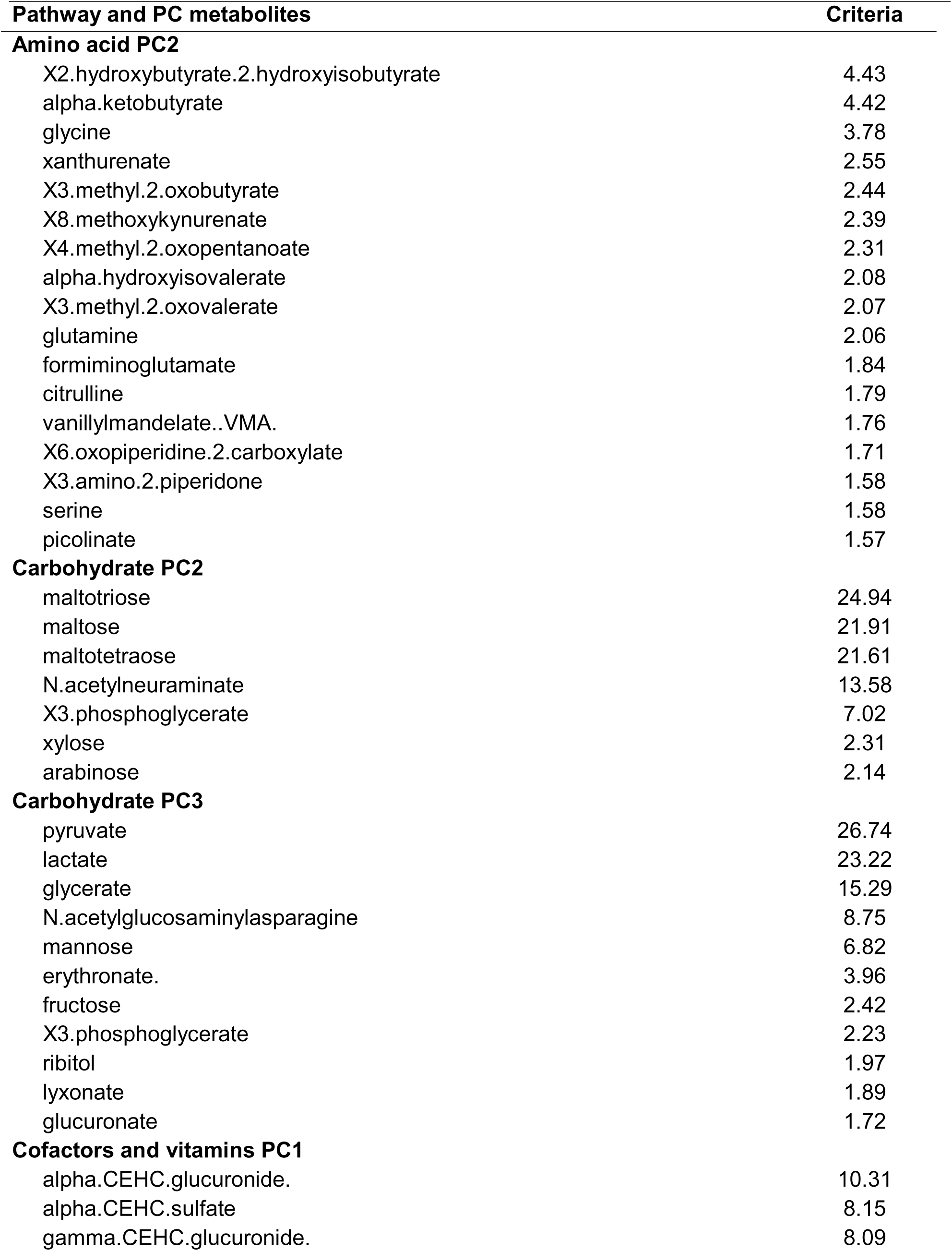
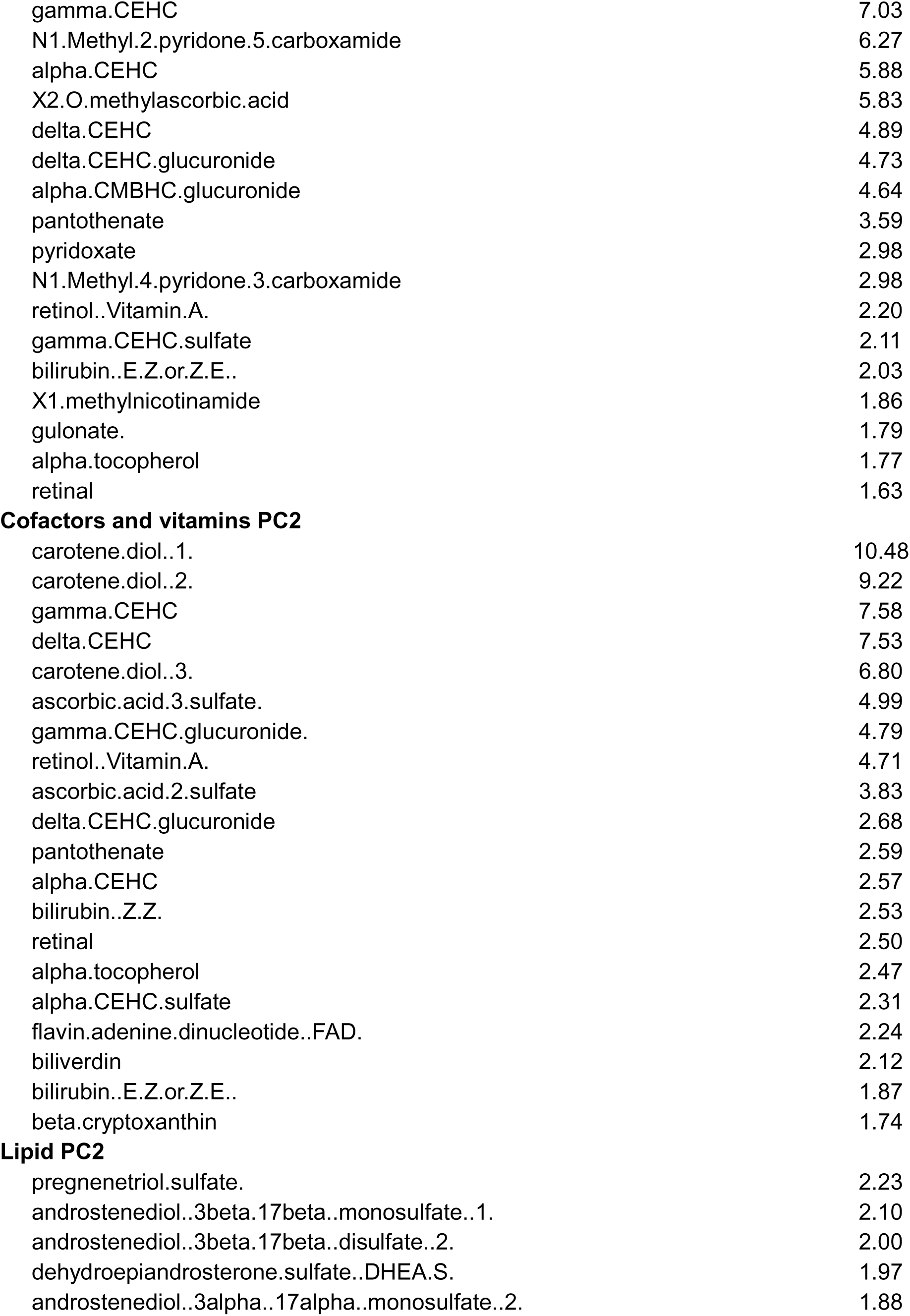
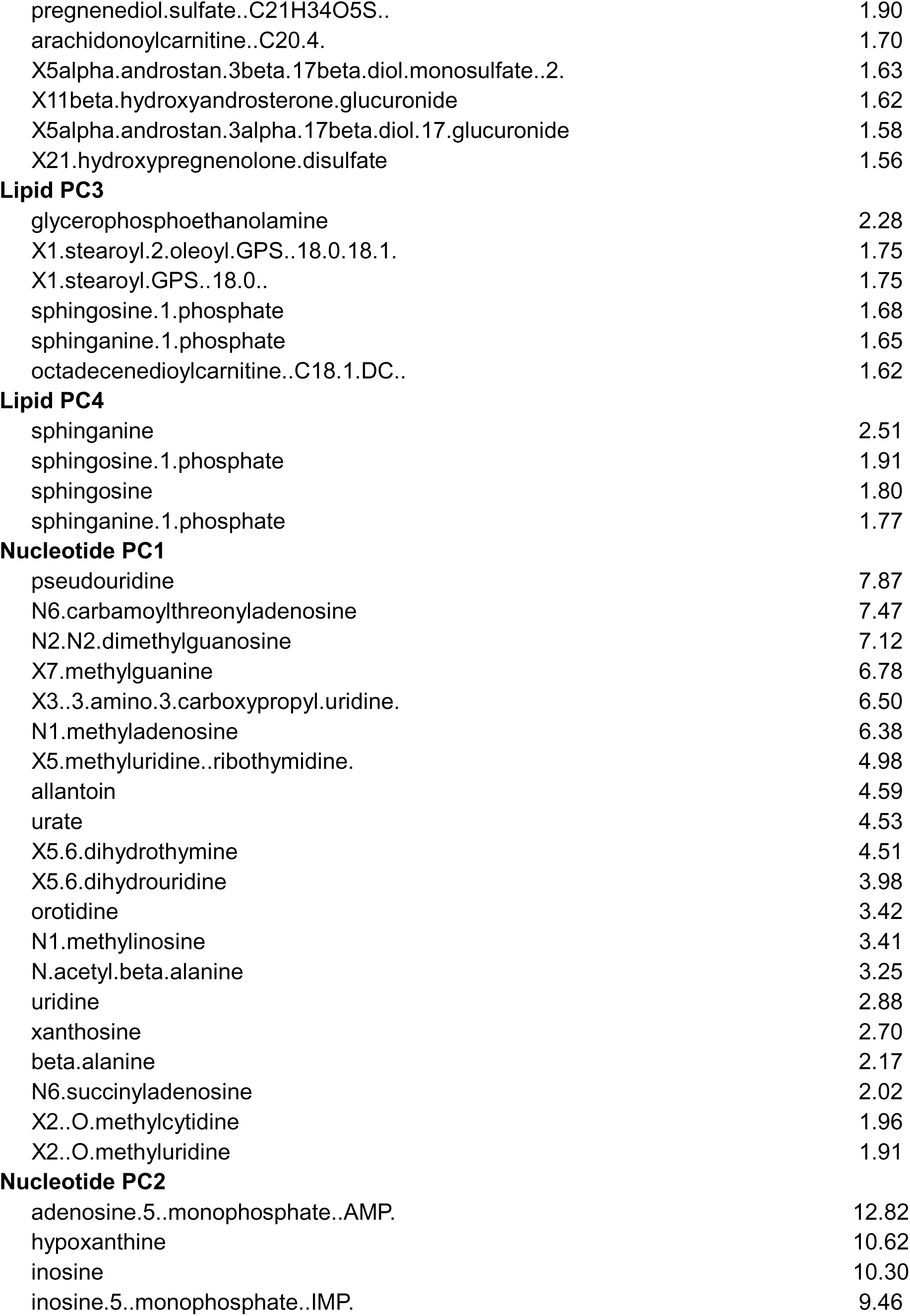
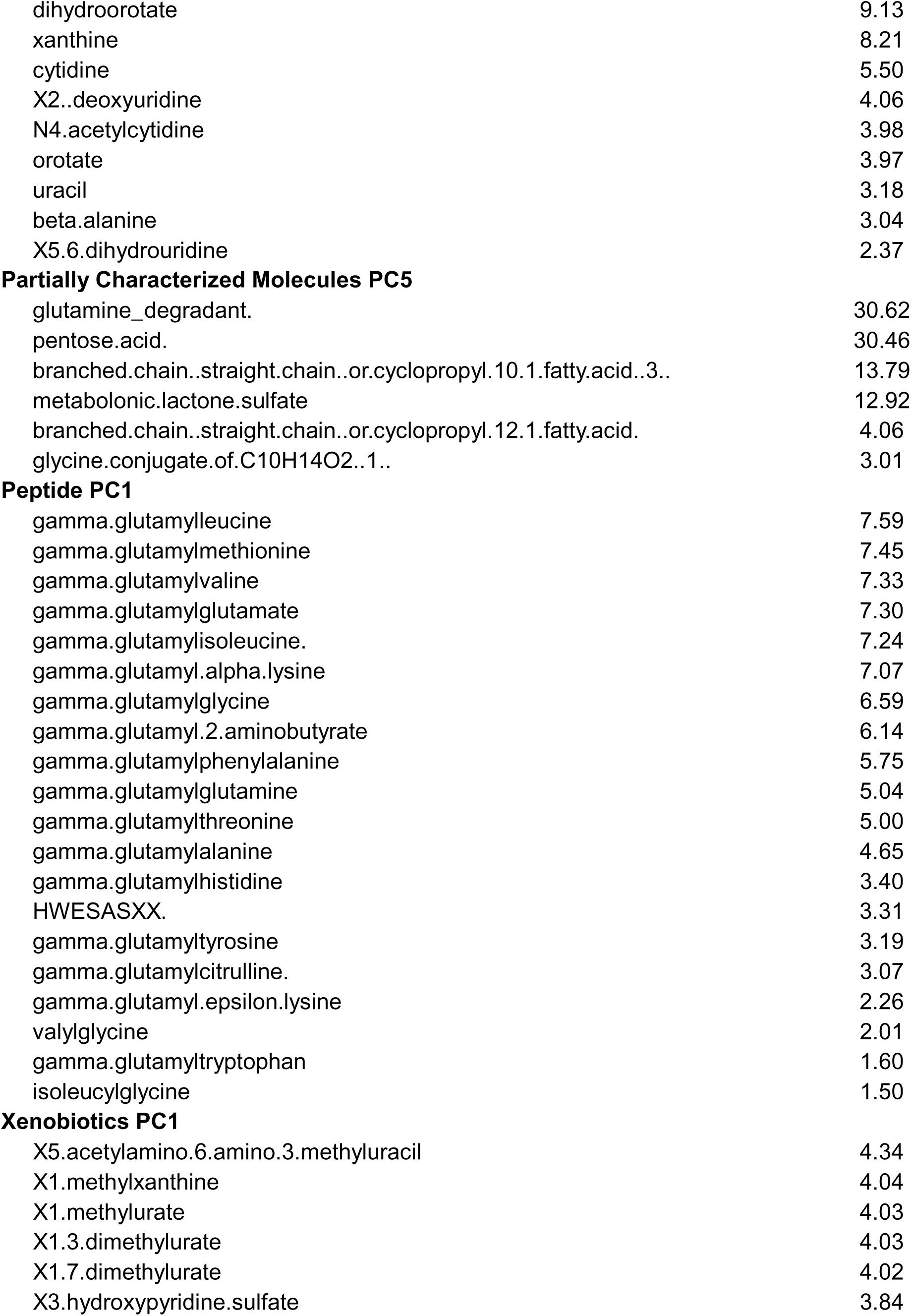
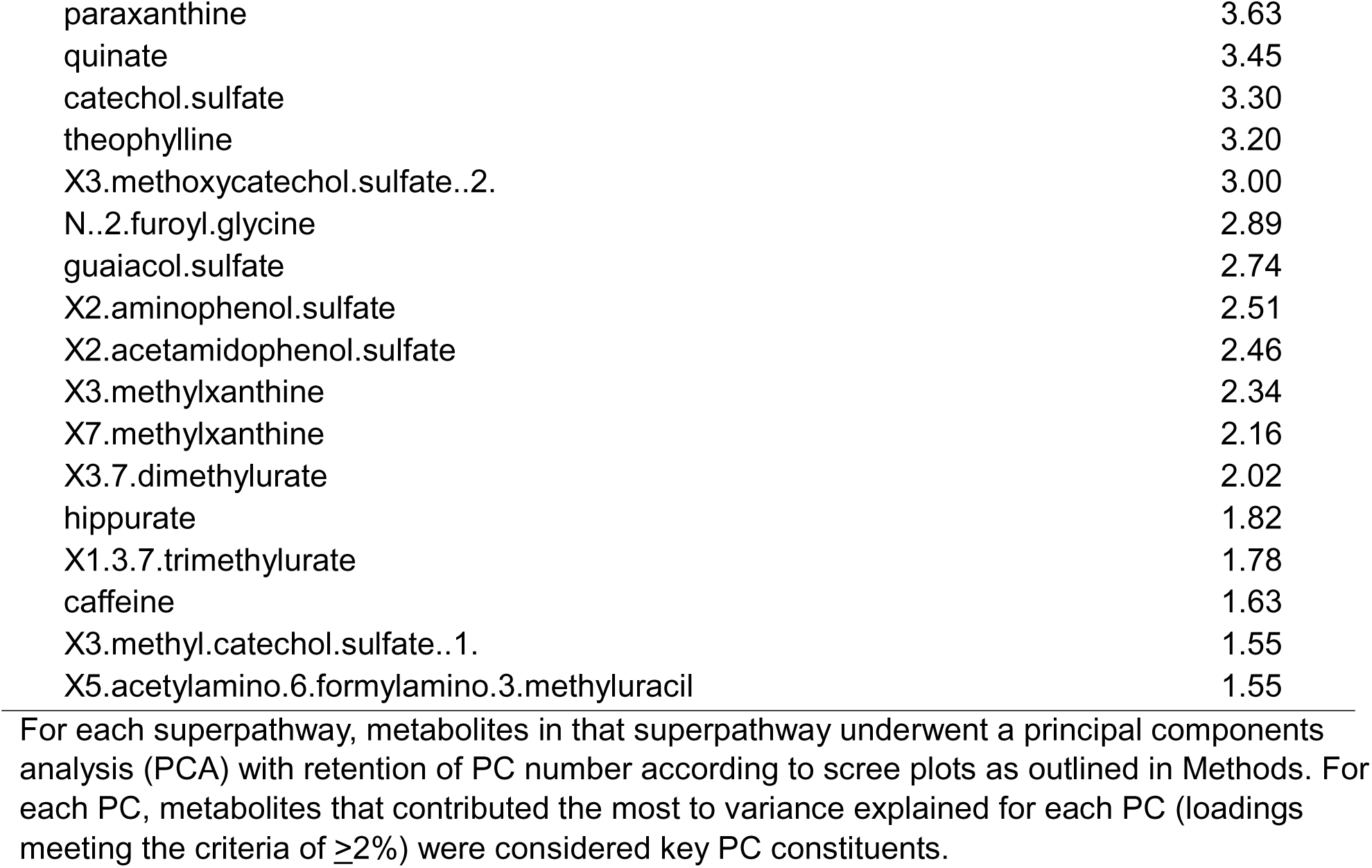
Metabolite contributions to Principal Components (PC).

